# The Geography of Excess Deaths in England during the Covid-19 pandemic: Longer term impacts and monthly dynamics

**DOI:** 10.1101/2020.09.07.20188003

**Authors:** Richard Breen, John Ermisch

## Abstract

Physical social interaction relevant to the spread of infectious diseases occurs, by its nature, at a local level. If infection and related mortality are associated with social background, it is therefore natural to study variation in them in relation to the social composition of local areas. The first part of the paper studies the geographical impact of Covid-19 infection on age-standardised sex-specific excess death rates during the peak months of the pandemic so far, March through May 2020. The second part examines monthly mortality dynamics in relation to predictions from a spatial SIR (Susceptible, Infected, Recovered) model of infection introduced by Bisin and Moro (2020). The analysis indicates that during the peak months of the Covid-19 pandemic, a larger non-white population and higher social deprivation in an area were associated with higher excess mortality, particularly among men. Regarding dynamics, higher population density accelerated the growth in mortality during the upsurge in infection and increased its rate of decline after the peak of the epidemic, thereby producing a more peaked mortality profile. There is also evidence of a slower post-peak decline in mortality in more socially deprived areas but a more rapid decline in areas with a larger non-white population.

## The Geography of Excess Deaths in England during the Covid-19 pandemic

Physical social interaction relevant to the spread of infectious diseases occurs, by its nature, at a local level. If infection and related mortality are associated with social background, it is therefore natural to study variation in them in relation to the social composition of local areas. Measurement of Covid-19 infection has been poor and variable across local areas because of inadequate testing for the virus. As a consequence, it is more reliable to study Covid-19 using mortality data based on death registrations rather than reports of confirmed cases, although of course Covid-19 related deaths follow some weeks after infection.

However, attribution of deaths to Covid-19 has been imperfect, even in the registration data, and so we construct a measure of ‘excess deaths’ by comparing deaths during the months of the Covid-19 pandemic with average deaths in the same months during previous years. The first part of the paper studies the impacts of Covid-19 infection using excess deaths during the peak months of the pandemic so far, March through May 2020. The second part examines monthly mortality dynamics in relation to predictions from a spatial SIR (Susceptible, Infected, Recovered) model of infection introduced by Bisin and Moro (2020).

We estimate that in England and Wales there were 55,219 excess deaths (standard error = 2,518) between March and May 2020.^1^ Our analyses indicate that during these peak months of the Covid-19 pandemic, higher social deprivation in an area and a larger non-white population were associated with higher excess mortality, particularly among men.

Regarding dynamics, higher population density accelerated the growth in mortality during the upsurge in infection and increased its rate of decline after the peak of the epidemic, thereby producing a more peaked mortality profile. There is also evidence of a slower post-peak decline in mortality in more socially deprived areas but a more rapid decline in areas with a larger non-white population.

### 1. Longer term excess mortality

The excess mortality measure used here is the age standardised mortality rate per 100,000 population (for each sex) for deaths occurring between 1st March and 31 May 2020 in English local authority areas (LA) minus the average age standardised mortality rate during 2014-2018 for that LA in the same part of the year.^2^ In calculating excess deaths the annual 2014-2018 average age standardised mortality rate in a LA was adjusted to March-May by applying the monthly fixed effects estimated from an analysis of data on monthly deaths since January 2006 (see Appendix Table 1). The monthly adjustment factors for March, April and May are 0.089, 0.084 and 0.08 of annual deaths, respectively, and so the three month March-May adjustment factor is 0.253. We focus on the period up to 31 May because, as we show below, excess mortality turned negative for four-fifths of the areas in June, making March through May the peak months of the pandemic in terms of mortality. Tables 1 and 2 show descriptive statistics for the 306 LA areas used in the analysis, and Figures 1 and 2 show the variation in excess death rates across English LAs.

**Table 1:** Age-standardised Mortality rates per 100,000, March to May 2020: summary statistics for all LAs

|  | Mean | Std. Dev. | Min | Max |
| --- | --- | --- | --- | --- |
| Male excess rate | 86.5 | 57.5 | -45.3 | 309.3 |
| Female excess rate | 56.5 | 37.1 | -26.2 | 163.7 |
| Male ave. 2014-18<sup>a</sup> | 279.3 | 34.5 | 137.2 | 385.9 |
| Female ave. 2014-18<sup>a</sup> | 207.2 | 25.7 | 109.2 | 279.3 |
| Male Covid-19 rate | 105.8 | 49.0 | 0.0 | 287.9 |
| Female Covid-19 rate | 61.2 | 28.5 | 0.0 | 150.7 |

<sup>a</sup>Annual average multiplied by 0.253.

**Table 2:**
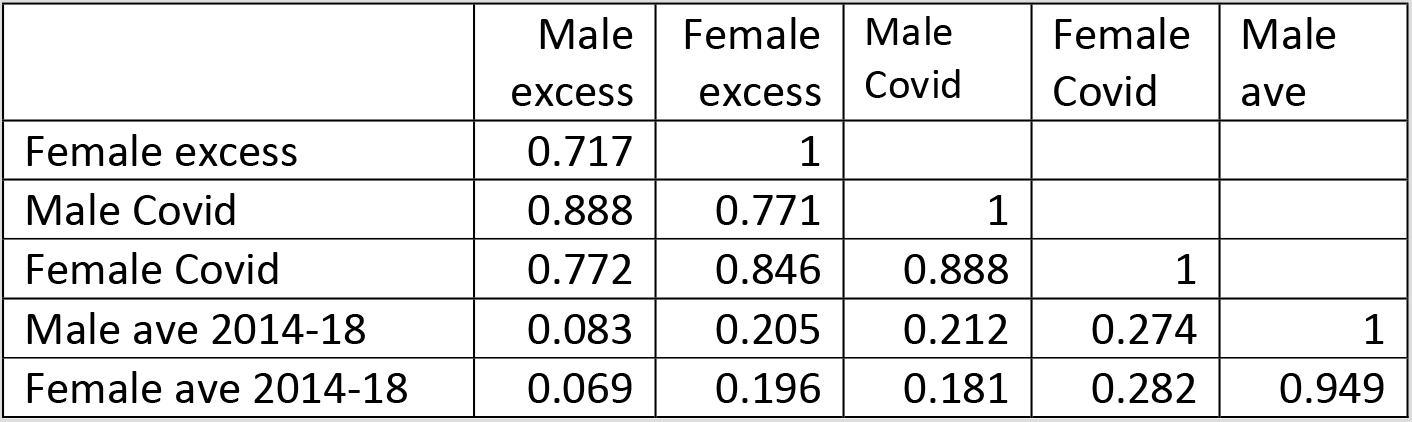
Age-standardised Mortality rates correlation matrix

**Figure 1:**
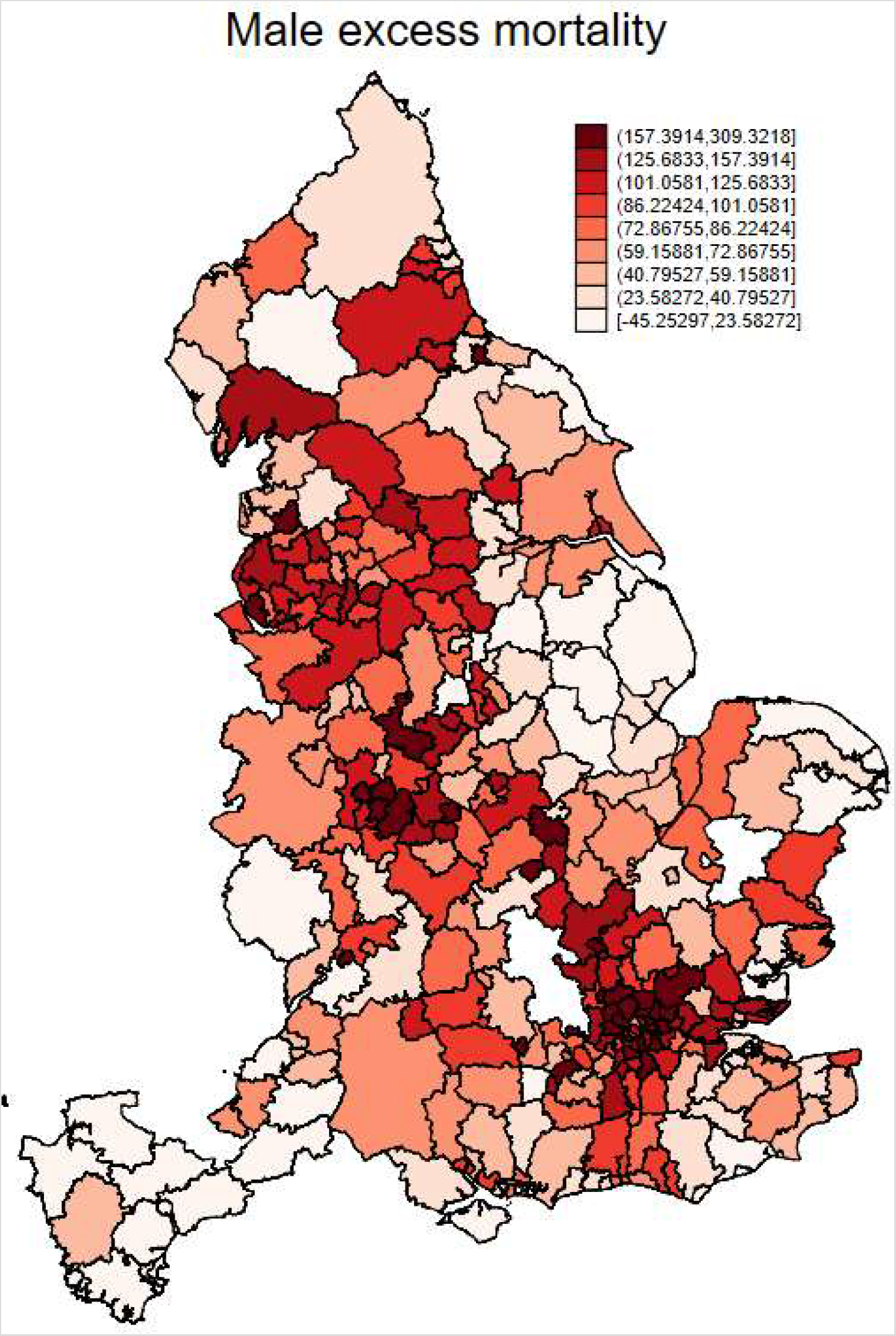
Male Excess Mortality per 100,000 population, 1 March to 31 May 2020

**Figure 2:**
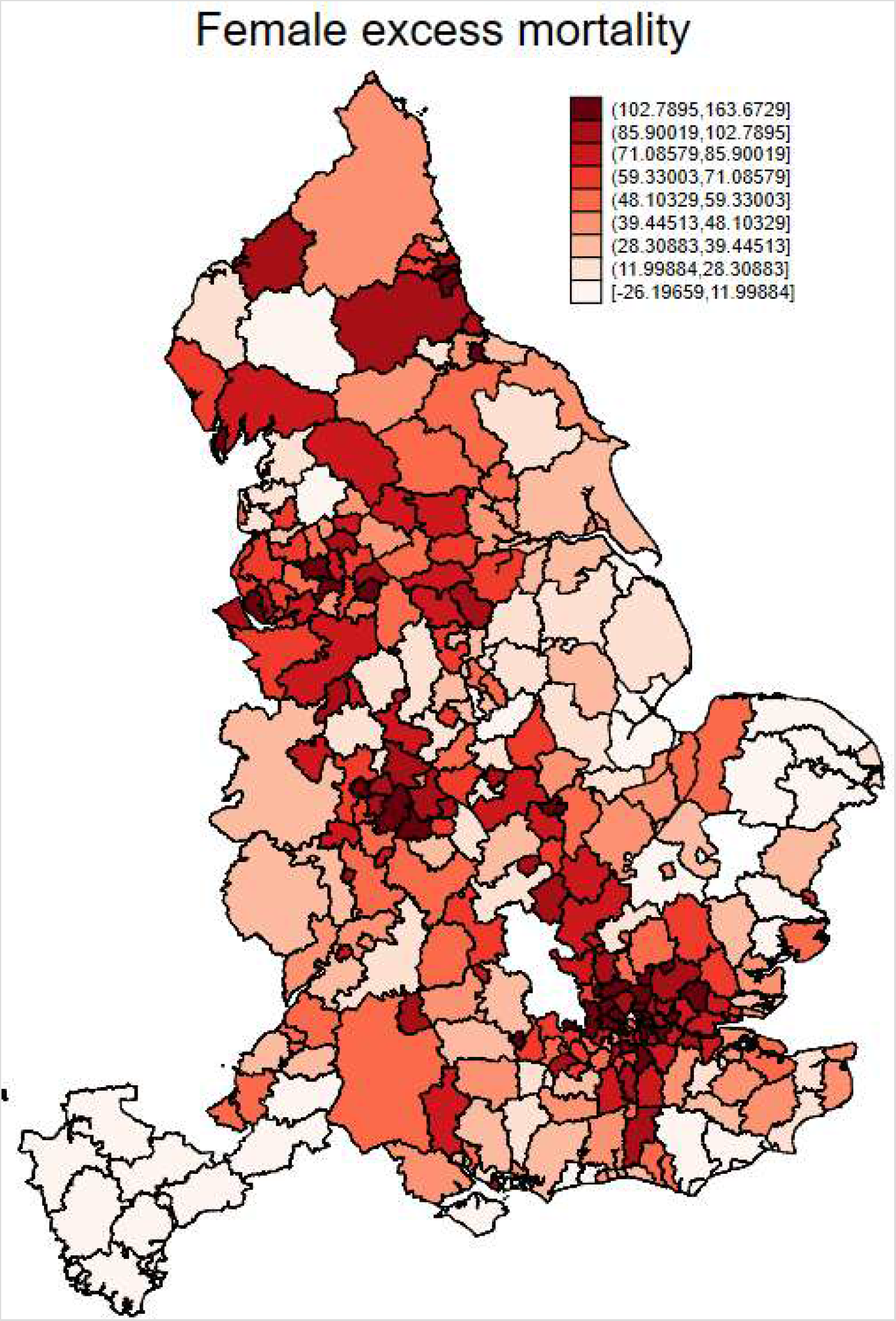
Female Excess Mortality per 100,000 population, 1 March to 31 May 2020

Table 1 shows women’s excess mortality is lower on average than men’s, and spatial variation in excess mortality is considerable for both sexes. The coefficient of variation across LAs is about 0.65 in each case, compared with 0.12 for average rates 2014-18, showing the greater geographical variability of excess deaths. The Covid-19 mortality rate is similarly higher for men than women; both have a coefficient of variation of 0.46.

Some of the highest excess mortality rates are in London boroughs. The three highest male rates are in Brent (258.9), Haringey (271.1) and Newham (309.3), while the lowest male rates are in rural areas, such as Torridge in North Devon (−45.3) and the Isle of Wight (−34.5). Very high female excess rates are not confined to London. For example, the highest is Watford (163.7) and the next highest is Middlesborough (156.6). Similar to males, the lowest female excess rates are in rural areas, with Devon highly represented among them, although the lowest is in Northeast Lincolnshire.

Table 2 shows a high correlation between Covid-19 mortality (as assessed from death registrations by ONS) and excess mortality (0.89 and 0.85 for men and women, respectively). Correlations between excess deaths and the longer run average rates are quite weak. Male and female Covid-19 death rates are more highly correlated (0.89) than between male and female excess death rates (0.72).

#### Socio-economic correlates

To measure the socio-economic composition of an area we use the Office of National Statistics’ (ONS) indices of an area’s relative deprivation, measured in 2019 (https://www.gov.uk/government/statistics/english-indices-of-deprivation-2019). Higher values indicate more deprivation in an area.^3^ We also consider ethnicity by including the proportion of the LA population who are non-white, based on population estimates by ONS for 2016.^4^ The mean proportion non-white in the sample is 0.11 (SD=0.13). Many, but not all, of the large values are in London: six non-London LAs have proportions larger than 0.30 (e.g. Leicester, Birmingham and Manchester) compared with 25 in London. In the spatial SIR model of Bisin and Moro (2020) population density plays a central role because it determines the number of contacts a person has on a given day and this drives the number of daily infections, along with the contagion rate per contact. Thus, we also include population density of the area as one of our predictors of excess mortality at the local level. Descriptive statistics for these variables are shown in Appendix Table 2.

#### Statistical Model for Spatial Variation in Excess Mortality

Because of evidence of spatial dependence^5^, our analysis uses a spatial regression model of the form:

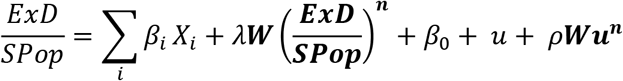

where 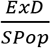is the age standardised excess mortality rate; the *Xs* are characteristics of areas; the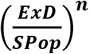 are excess death rates in neighbouring areas; **W** is a spatial weighting matrix based on contiguity of areas; *λ* is a spatial spill-over parameter; the disturbance *u* also reflects errors in measuring the mortality rate at the local area level and **u^n^** are residual influences on mortality in neighbouring areas. In practice it is often difficult to estimate both *λ* and *ρ* with precision, and we will focus on a model with *λ* only because the estimates of *β*_i_ are virtually unaffected by also allowing for residual spatial correlation.^6^ Despite providing this structure, the analysis makes no causal claims, and *β_i_* only reflects spatial associations between area social composition and excess deaths. Estimation of the model is carried out by generalised method of moments using the spregress commands in Stata 15.1.

The parameter estimates in the first two columns of Table 3 indicate that both male and female excess mortality are associated with an area’s social deprivation score. To provide a guide to magnitudes, a ten percent higher social deprivation index is associated with 1.2 per 100,000 higher excess mortality among men (1.0 per 100,000 among women).

**Table 3:** Estimation results: Excess and Covid-19 Mortality per 100,000, 1 March to 31 May 2020 (N=306)

| Parameter | Males Ex.<br>(se) | Females Ex.<br>(se) | Males Covid<br>(se) | Females Covid<br>(se) |
| --- | --- | --- | --- | --- |
| $\beta \ln(\text{ave. score})$ | 12.4<br>(6.0) | 10.1<br>(4.4) | 16.3<br>(4.0) | 12.0<br>(3.1) |
| $\beta$ Proportion non-white | 159.8<br>(24.9) | 68.7<br>(17.8) | 144.8<br>(21.2) | 60.0<br>(13.3) |
| $\beta \ln(\text{pop. density})$ | 6.7<br>(2.3) | 6.6<br>(1.8) | 9.6<br>(2.0) | 6.1<br>(1.3) |
| $\lambda W \ln\left(\frac{D}{Pop}\right)^n$ | 0.46<br>(0.07) | 0.36<br>(0.08) | 0.31<br>(0.05) | 0.33<br>(0.05) |
| *Intercept* | -44.3<br>(20.4) | -40.2<br>(15.2) | -47.2<br>(16.0) | -36.9<br>(10.9) |
| Pseudo R<sup>2</sup> | 0.530 | 0.364 | 0.636 | 0.509 |
| Wald test of spatial terms, p-value | <0.0001 | <0.0001 | <0.0001 | <0.0001 |

se refers to the robust standard error allowing for heteroskedasticity.

However, while male and female excess deaths are also greater the larger the proportion non-white in an area, this relationship is much stronger among men A 0.01 higher proportion non-white is associated with of 1.6 per 100,000 higher excess mortality for men compared with 0.7 per 100,000 for women. Population density also has a significant effect. A ten percent increase in area population density increases excess mortality by about 0.7 per 100,000. The estimates of *λ* indicate a relatively strong spillover of excess mortality from contiguous areas.

The last two columns of Table 3 show comparable parameter estimates for Covid-19 mortality. The patterns of association with area social deprivation, proportion non-white and population density are similar to those for excess mortality. The proportion of the spatial variance of Covid-19 mortality accounted for by these variables is larger than for excess mortality. Comparison with non-Covid-19 mortality in the same period (see Appendix Table 3) indicates that the patterns in relation to social composition are very different: non-Covid-19 mortality is much more strongly associated with social deprivation than Covid-19 mortality, and non-Covid-19 mortality is negatively associated with the proportion non-white in the LA. Non-Covid-19 mortality is also associated less with population density and with mortality in contiguous areas.

#### Monthly excess mortality

Table 4 reports summary statistics for the individual months of April, May and June 2020, from which it is clear that excess deaths were much lower in May and negative for the vast majority of LAs in June (86% for men, 79% for women). The English and Welsh national figures shown in Figure 3 indicate that excess mortality in July continued to be low: during the period 4 July to 31 July, excess deaths in England and Wales were negative, representing 2.5% of average deaths during 2015-2019. It was only in the second week of August that excess deaths became positive again, continuing so through the third week.

**Table 4:**
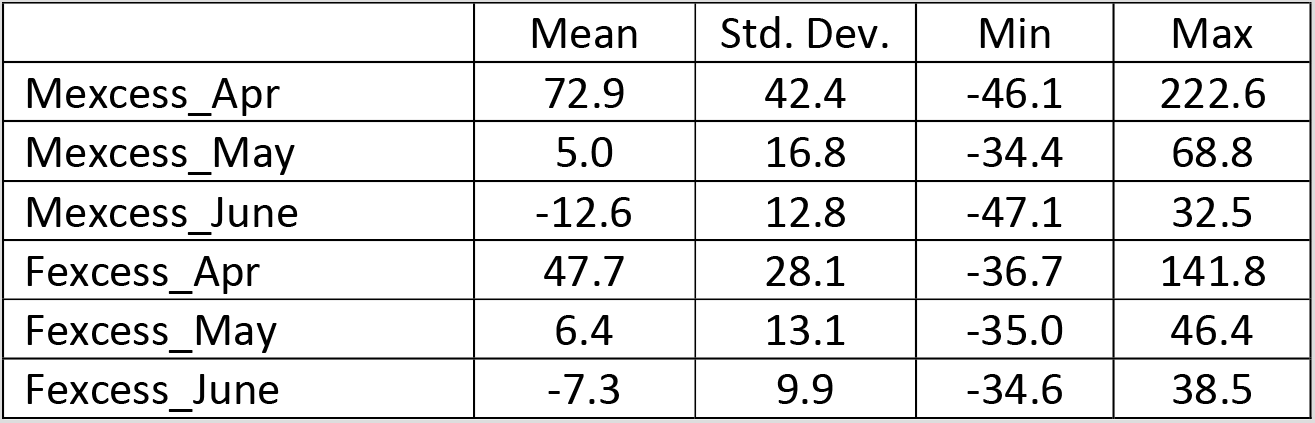
Age-standardised Mortality rates per 100,000 in local areas: summary statistics

**Figure 3:**
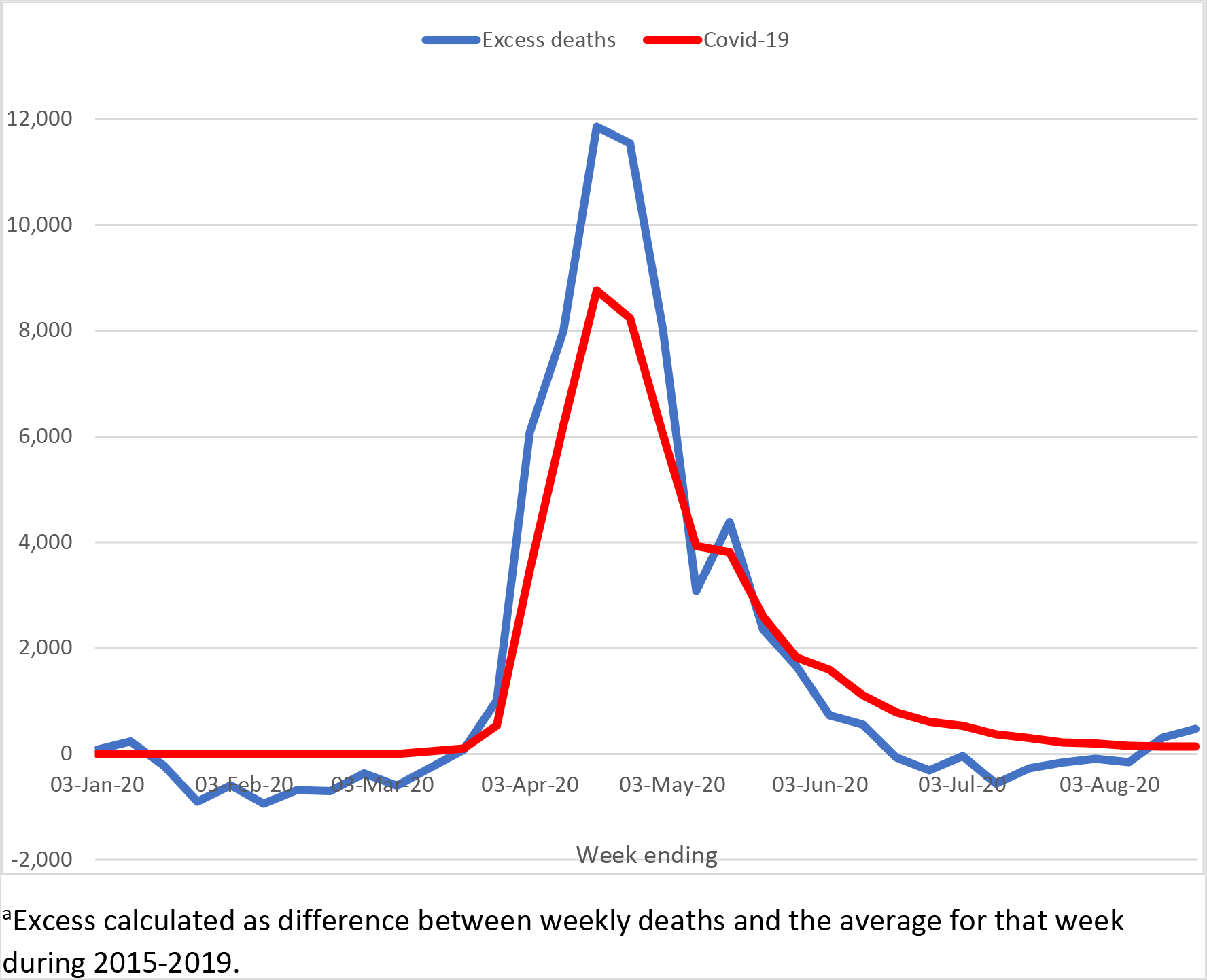
Weekly excess deaths and Covid-19 deaths, England and Wales^a^

Source: https://www.ons.gov.uk/peoplepopulationandcommunity/birthsdeathsandmarriages/death s/datasets/weeklyprovisionalfiguresondeathsregisteredinenglandandwales

Table 5 shows that among both men and women the associations between the area social composition variables are only present to any important degree in April, during the peak weeks of the pandemic, not in May or June. Indeed, the only serious influence on an area’s excess mortality during May was excess mortality in contiguous local authorities, as indicated by the high estimate of *λ* in May, and even that influence did not operate in June.

**Table 5:** Estimation results: Monthly Excess Mortality per 100,000 (N=306)

| Parameter | Males,<br>April<br>(se) | Males,<br>May<br>(se) | Males,<br>June<br>(se) | Females,<br>April<br>(se) | Females,<br>May<br>(se) | Females,<br>June<br>(se) |
| --- | --- | --- | --- | --- | --- | --- |
| $\beta \ln(\text{ave. score})$ | 9.3<br>(4.3) | 3.9<br>(2.7) | -0.9<br>(1.8) | 4.1<br>(3.3) | 4.7<br>(2.0) | 1.0<br>(1.7) |
| $\beta$ Proportion non-white | 117.9<br>(22.6) | -2.0<br>(9.0) | -1.5<br>(6.7) | 55.9<br>(15.3) | -10.1<br>(7.1) | -12.0<br>(6.4) |
| $\beta \ln(\text{pop. density})$ | 6.7<br>(1.9) | -0.1<br>(1.0) | -1.0<br>(0.8) | 5.6<br>(1.4) | 0.5<br>(0.9) | -0.5<br>(0.6) |
| $\lambda W \ln\left(\frac{D}{Pop}\right)^n$ | 0.38<br>(0.07) | 0.98<br>(0.23) | -0.02<br>(0.2) | 0.34<br>(0.08) | 0.66<br>(0.19) | 0.01<br>(0.3) |
| *Intercept* | -33.4<br>(16.8) | -10.1<br>(8.2) | -3.1<br>(6.6) | -20.0<br>(12.0) | -12.8<br>(5.9) | 05.2<br>(5.9) |
| Pseudo R<sup>2</sup> | 0.525 | 0.040 | 0.021 | 0.389 | 0.058 | 0.046 |
| Wald test of spatial terms, p-value | <0.0001 | <0.0001 | 0.93 | <0.0001 | 0.0005 | 0.963 |

se refers to the robust standard error allowing for heteroskedasticity.

In models not reported, the April excess mortality rate was included in the equation for May excess mortality, and analogously for June excess mortality. These indicated neither persistence nor reversal over time.

The story about excess deaths in relation to area social composition is, therefore, a story about what happened during the peak of the pandemic. During this period, a larger non-white population and higher social deprivation in an area were associated with higher excess mortality, the former having a particuarly strong relationship with male excess mortality.

### 2. Monthly mortality dynamics

According to the spatial SIR model of Bisin and Moro (2020), the steady-state fraction infected (i.e. (recoveries + deaths)/population) increases with the population density of an area. The model also predicts that higher density increases the slope of the curve of active cases per head of population (*I*/*N*) and increases its peak, as illustrated in Figure 4.

**Figure 4:**
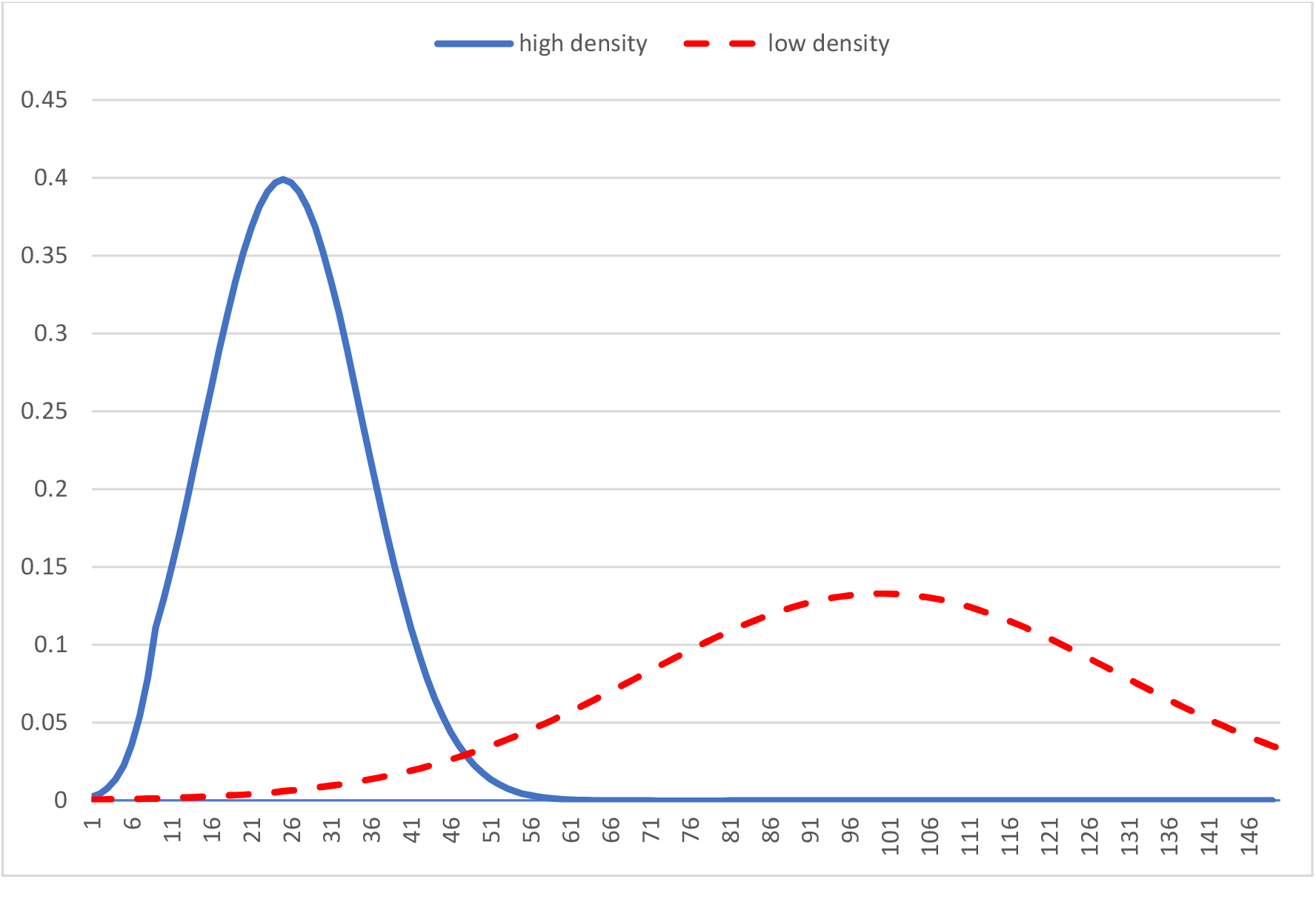
Active cases per head (*I*/*N*) by day of the epidemic, high and low density areas high density low density

Source: Stylised curves suggested by simulation on the right side of Figure 10 in Bisin and Moro (2020).

One of the model’s prediction is that the growth rate of the fraction infected, 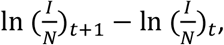 other things equal, is proportional to the density of the area. We examine this proposition with monthly mortality data from local authorities in England and Wales during March to June 2020.

Let Covid-19 deaths be proportional to active cases (*CD* = *δI*) so that, 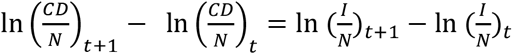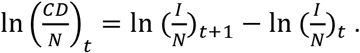 Deaths can be decomposed into ‘normal’ deaths and excess deaths due to the epidemic. Taking any two months during epidemic, the log change in the total death rate 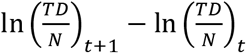 is made up of the normal increase in deaths between these two months and excess deaths due to Covid-19. Density should not affect the normal log change between months, but it does affect the change in excess deaths according to the Bisin-Moro model, and the model predicts that higher density increases the rate of increase in mortality up to the peak and increases the rate of decline (‘more negative’) after the peak, leading to a sharper peak in more dense areas.

In England and Wales, the peak of the epidemic was in April, leading us to expect that density would have a positive impact on the log change in mortality rate from March to April and a negative impact from April to May. By June, estimates of excess mortality are negative in most areas, and so we should no longer expect density to affect the log change in mortality.

In our estimates we also allow for the area infection rate to vary with its social deprivation (e.g. people with lower level jobs may have to have more contact with others and there may be proportionately more of them in socially deprived areas) and with the proportion non-white.^7^ As before, we focus on the age standardised mortality rate for each sex, and the analysis of its log change over each two month interval is carried out using a spatial autoregression model with a ‘spatially lagged’ dependent variable and a spatial weighting matrix based on contiguity of areas.

The parameter estimates for density in Table 6 are consistent with the predictions of the Bisin-Moro model: higher density increases the slope of mortality during the upsurge (March to April) and reduces it after the peak, thereby producing a more peaked mortality profile. There is also evidence of a slower post-peak decline in mortality in more socially deprived areas, and a sharper decline in areas with a larger non-white population. Similar results obtain for female mortality.

**Table 6:** Estimation results: log change in Male Mortality per 100,000 (N=306)

| Parameter | March- April<br>(se) | April-May<br>(se) | May-Jun<br>(se) |
| --- | --- | --- | --- |
| $\beta \ln(\text{pop. density})$ | 0.05<br>(0.01) | -0.08<br>(0.01) | 0.00<br>(0.01) |
| $\beta \ln(\text{ave SD score})$ | 0.00<br>(0.03) | 0.08<br>(0.04) | -0.02<br>(0.04) |
| $\beta \text{ Proportion non-white}$ | 0.01<br>(0.10) | -0.62<br>(0.14) | 0.03<br>(0.12) |
| $\lambda (W(\ln(TD/N)_{t+1} - \ln(TD/N)_t))$ | 0.35<br>(0.08) | 0.35<br>(0.07) | 0.33<br>(0.16) |
| *Intercept* | -0.02<br>(0.12) | -0.06<br>(0.14) | -0.11<br>(0.13) |
| Pseudo R<sup>2</sup> | 0.139 | 0.447 | 0.012 |
| Wald test of spatial terms, p-value | <0.0001 | <0.0001 | <0.0001 |

se refers to the robust standard error allowing for heteroskedasticity.
$W$ is a spatial weighting matrix based on contiguity of areas; $\lambda$ is a spatial spill-over parameter

## Data Availability

All data are publicly available from the UK Office of National Statistics, and links are provided within the paper and in next box below.

https://www.ons.gov.uk/releases/deathsinvolvingcovid19bylocalareaandsocioeconomicdeprivationdeathsoccurringbetween1marchand30june2020

https://www.ons.gov.uk/peoplepopulationandcommunity/birthsdeathsandmarriages/deaths/datasets/deathsregisteredbyareaofusualresidenceenglandandwales.

https://www.gov.uk/government/statistics/english-indices-of-deprivation-2019

https://www.ons.gov.uk/peoplepopulationandcommunity/populationandmigration/populationestimates/datasets/populationcharacteristicsresearchtables

## Appendix 1

The analysis of monthly death counts in England and Wales allows for a cubic trend over the 175 months from January 2006 (Date=1) through July 2020 (Date=175) and for fixed effects for each month of the year (April being the reference month, the average deaths in which are indicated by the equation’s constant). Excess deaths are estimated from individual dummy variables for March, April, May, June and July 2020, the months of the Covid-19 epidemic included in the available monthly data.

**Appendix Table 1:** Regression estimates of Monthly Deaths January 2006-July 2020 (robust standard error allowing for heteroskedasticity below coefficient)

| Month: | Par.<br>Est. <sup>a</sup> | Adj.<br>factor<sup>b</sup> |
| --- | --- | --- |
| January | 9,923 | 0.103 |
| February | 1,008 | 0.086 |
| March | 2,686 | 0.089 |
| May | -1,882 | 0.084 |
| June | -3,529 | 0.080 |
| July | -3,615 | 0.077 |
| August | -5,161 | 0.077 |
| September | -4,374 | 0.074 |
| October | -1,024 | 0.076 |
| November | -690 | 0.082 |
| December | 2,446 | 0.083 |
| Eqn.constant<br>(April deaths) | 43,323 | 0.084 |
| Cubic Trend: |  |  |
| Date | -102<br>(40) |  |
| Date-sq | 1.461<br>(0.543) |  |
| Date-cu | -0.005<br>(0.002) |  |
| Mar-2020 | **2,238**<br>**(1,001)** |  |
| Apr-2020 | **43,457**<br>**(1,117)** |  |
| May-2020 | **9,524**<br>**(944)** |  |
| June-2020 | **1,459**<br>**(974)** |  |
| July-2020 | **-273**<br>**(934)** |  |
| R<sup>2</sup> | 0.830 |  |
| DW | 1.98 |  |
| Durbin alt: p-value |  |  |
| Lag 1 | 0.977 |  |
| Lag 2 | 0.970 |  |

<sup>a</sup>Standard errors of monthly fixed effects range from 1,004-1,510.
<sup>b</sup>Monthly adjustment factor is average monthly deaths divided by annual deaths

Summing the parameter estimates for the months March to July 2020 provides our estimate of 56,405 excess deaths during this period (robust SE=2,628), and analoglously 55,219 excess deaths (robust SE=2,518) during March to May 2020.

**Appendix Table 2:** Descriptive statistics (N=306)

|  | Mean | Std. Dev. | Min | Max |
| --- | --- | --- | --- | --- |
| ln(ave. score) | 2.901 | 0.421 | 1.713 | 3.808 |
| Proportion non-white | 0.110 | 0.135 | 0 | .0.684 |
| ln(pop. density) | 6.635 | 1.399 | 3.219 | 9.686 |

**Appendix Table 3:** Non-Covid-19 mortality, March-May 2020 (N=306)

| Parameter | Males Ex.<br>(se) | Females Ex.<br>(se) |
| --- | --- | --- |
| $\beta \ln(\text{ave. score})$ | 61.5<br>(4.7) | 47.0<br>(3.6) |
| $\beta$ Proportion non-white | -48.1<br>(20.2) | -53.9<br>(16.9) |
| $\beta \ln(\text{pop. density})$ | 3.1<br>(2.2) | 3.4<br>(1.9) |
| $\lambda W \ln\left(\frac{D}{Pop}\right)^n$ | 0.08<br>(0.02) | 0.04<br>(0.02) |
| *Intercept* | 53.8<br>(17.5) | 46.3<br>(14.1) |
| Pseudo R<sup>2</sup> | 0.387 | 0.366 |
| Wald test of spatial terms, p-value | 0.0005 | 0.0678 |

se refers to the robust standard error allowing for heteroskedasticity.

## Footnotes

1 Estimate based on a statistical model of monthly death counts for England and Wales using monthly data from January 2006 to July 2020, outlined in the Appendix. Between March and July 2020 there were 56,405 excess deaths (standard error = 2,628), indicating that most of the excess deaths in 2020 through July were during March to May.

2 Age-standardised mortality rates for each sex are standardised to the 2013 European Standard Population. They allow for differences in the age structure of populations and therefore allow valid comparisons to be made between geographical areas, the sexes and over time. Table 1 of the UK Office of National Statistics (ONS) publication Deaths involving COVID-19 by local area and deprivation: deaths occurring between 1 March and 30 June 2020 in England and Wales (24 July 2020) indicates the confidence intervals around the estimates https://www.ons.gov.uk/releases/deathsinvolvingcovid19bylocalareaandsocioeconomicdepri vationdeathsoccurringbetween1marchand30june2020 The source for age standardised death rates by LA during 2014 through 2018 is: https://www.ons.gov.uk/peoplepopulationandcommunity/birthsdeathsandmarriages/deaths /datasets/deathsregisteredbyareaofusualresidenceenglandandwales.

3 The analysis considered the overall index as well as its health and income components. The three measures are highly correlated, and analyses using the health and income components yielded no new insights concerning mortality.

4 https://www.ons.gov.uk/peoplepopulationandcommunity/populationandmigration/populati onestimates/datasets/populationcharacteristicsresearchtables.

5 The Moran test for spatial dependence produces p-values < 0.0001 for all the mortality variables analysed.

6 In both male and female equations, the estimates of the *β*_*i*_ parameters are very similar in the specification with only the ‘spatially lagged dependent variable’ to a model with both it and residual spatial autocorrelation, and the estimate of *ρ* is imprecisely estimated (smaller than or about the same as its standard error).

7 Although relating to Covid-19 deaths rather than infections, ONS mortality data up to 20 April is suggestive. It indicates that ‘compared with the rate among people of the same sex and age in England and Wales, men working in the lowest skilled occupations had the highest rate of death involving Covid-19…’. Also, ‘men and women working in social care, a group including care workers and home carers, both had significantly raised rates of death involving Covid-19…’ See https://www.ons.gov.uk/peoplepopulationandcommunity/healthandsocialcare/causesofdeath/bulletins/coronaviruscovid19relateddeathsbyoccupationenglandandwales/deathsregistereduptoandincluding20april2020 To the extent that non-white people are overly represented in such occupations, an analogous argument may apply to the proportion non-white in an area.

